# Longitudinal variation in SARS-CoV-2 antibody levels and emergence of viral variants: implications for the ability of serological assays to predict immunity

**DOI:** 10.1101/2021.07.02.21259939

**Authors:** Frauke Muecksch, Helen Wise, Kate Templeton, Becky Batchelor, Maria Squires, Kirsty McCance, Lisa Jarvis, Kristen Malloy, Elizabeth Furrie, Claire Richardson, Jacqueline MacGuire, Ian Godber, Alana Burns, Sally Mavin, Fengwen Zhang, Fabian Schmidt, Paul Bieniasz, Sara Jenks, Theodora Hatziioannou

## Abstract

**Background:** Serological assays are being deployed to monitor antibody responses in SARS-CoV-2 convalescents and vaccine recipients. There is a need to determine whether such assays can predict immunity, as antibody levels wane and viral variants emerge.

**Methods:** We measured antibodies in a cohort of SARS-CoV-2 infected patients using several high-throughput serological tests and functional neutralization assays. The effects of time and spike protein sequence variation on the performance and predictive value of the various assays was assessed.

**Findings:** Neutralizing antibody titers decreased over the first few months post-infection but stabilized thereafter, at about 30% of the level observed shortly after infection. Serological assays commonly used to measure antibodies against SARS-CoV-2 displayed a range of sensitivities that declined to varying extents over time. Quantitative measurements generated by serological assays based on the spike protein were better at predicting neutralizing antibody titers than assays based on nucleocapsid, but performance was variable and manufacturer positivity thresholds were not able to predict the presence or absence of detectable neutralizing activity. Even though there was some deterioration in correlation between serological measurements and functional neutralization activity, some assays maintained an ability to predict neutralizing titers, even against variants of concern.

**Interpretation:** The ability of high throughput serological assays to predict neutralizing antibody titers is likely crucial for evaluation of immunity at the population scale. These data will facilitate the selection of the most suitable assays as surrogates of functional neutralizing activity and suggest that such measurements may have utility in clinical practice.

## Introduction

The world has experienced an unprecedented pandemic following the emergence of severe acute respiratory coronavirus 2 (SARS-CoV-2). Millions have died and the repercussions have affected every aspect of life. The remarkable mobilization of the scientific community in response to the pandemic has led to the rapid development of safe and effective vaccines, as well as reagents and assays to aid in the detection and mitigation of virus spread.

An early prominent issue in the pandemic was the accurate identification of infected individuals at a large scale. While PCR-based assays remain a reliable and sensitive test for infection they are not amenable to mass population screening. Thus, serological assays, despite limitations,^1-3^ have been instrumental for surveillance and providing selection criteria to recruit vaccine trial participants and convalescent plasma donors. Monitoring antibody titers is necessary to measure the magnitude and longevity of immune responses induced by natural infection or vaccination. As immune responses to SARS-CoV-2 antigens are increasingly elicited by infection and/or vaccination, the measurement of antibody titers and the ability of such measurements to predict protection from infection or disease will be of great importance. Whether simple serological tests will be able to predict neutralizing antibody titers and immunity to SARS-CoV-2 is yet to be determined. Moreover, as antibodies both mature, acquiring greater affinity, while total levels decline, ^4-6^ and new SARS-CoV-2 variants emerge, the predictive value of serological tests based on the prototype viral strain will need to be evaluated.

A number of high-throughput serological assays are routinely used to detect antibodies against nucleocapsid (N) or spike (S) viral antigens. These assays were initially designed to provide a positive or negative test result, but they also generate quantitative measurements of antibody levels. Prior studies of how these quantitative serological values correlate with neutralizing antibodies titers have yielded variable results^7-9^. Importantly, the sensitivity of the assays as diagnostic tools and their predictive value for immune parameters several months after infection and against variants of concern has not been assessed. We present a longitudinal study of COVID-19 recovered patients over 6 months post-infection, evaluating the diagnostic sensitivity of ten different serological assays and their ability to predict neutralizing antibody activity against SARS-CoV-2.

## Results

A previously reported^8^ cohort of participants that developed mild symptoms following SARS-CoV-2 infection, was repeatedly sampled, up to 7.2 months post-infection, to evaluate how neutralizing antibody levels correlate over time with antibodies measured using ten high throughput serological assays. Neutralizing antibody titers were measured at up to five visits for each participant using a pseudovirus neutralization assay that correlates well with neutralization against authentic SARS-CoV-2^10^. Consistent with prior studies^4,8,11,12^, the half-maximal neutralizing titers (NT50) declined over time in the majority of patients (Figure 1A,B). The most significant rate of decrease, approximately 25%, was observed between the early visits reaching a 45% decrease in NT50 by visit 3, approximately 70 days post-infection (Figure 1C). Thereafter the rate of decrease became less pronounced and NT50 values at visits 4 and 5, at approximately 3-7 months post-infection, appeared to stabilize at ∼30% of the levels observed within the first 2 months post-infection (Figure 1B,C). NT50 values were higher in male than female participants at visit 1, but the overall rate of NT50 decline from the first to the last visit was greater for male participants (supplementary Figure 1). Thus, the sex difference gradually diminished and was not discernable by visit 4. No correlation between NT50 values and age were observed at any time point (supplementary Figure 1).

**Figure 1.**
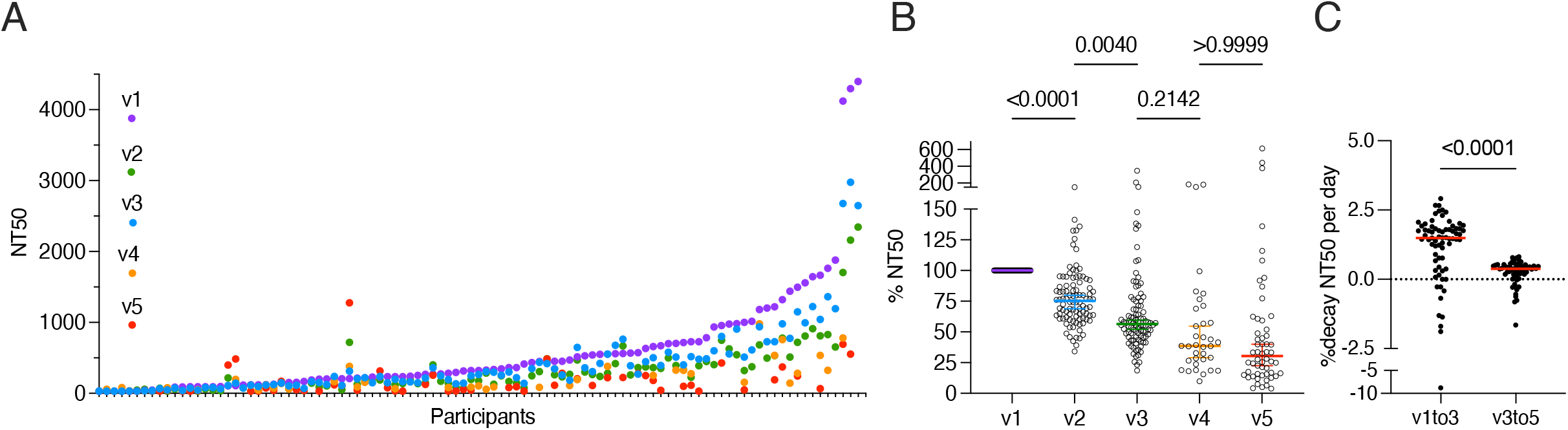
Neutralization activity in longitudinal COVID-19 patient sera. **(A)** Half-maximal neutralization titers (NT50s) for each sample collected at the visit indicated (v1-5). **(B)** Relative NT50 values in sera obtained at visit 1-5, normalized to visit 1. Colored horizontal bars indicate median values with 95% confidence interval. Statistical significance was determined with Kruskal-Wallis test and Dunn’s multiple comparisons test. **(C)** Relative decay of NT50 per day between visits. Red horizontal bars indicate median. Statistical significance was assessed with the Wilcoxon test.

The same sera were analyzed using nine different serological assays that detect antibodies against either the viral nucleocapsid (N) protein, or various forms of the spike (S) protein that included trimeric S, S1/S2 subunits or the receptor binding domain (RBD). First, the sensitivity of each assay was determined for three time windows over the course of the study. All assays were sensitive at the first window, 21-80 days post-infection; Abbott IgGII Quant, Roche S and Roche N had the highest sensitivities at 100%, Siemens COV2T and Diasorin Trimeric S were 95% sensitive and Diasorin S1/S2, Euroimmun and Abbott (N) ranged from 85-90%. While Abbott IgGII Quant, Roche N and Diasorin S1/S2 maintained their sensitivity over time, the sensitivity of other assays declined to varying degrees, ranging from 45% to 85% at >140 days post-infection (Figure 2A). Thus, the performance of the assays for serosurveillance applications at >140 days after infection was extremely variable.

**Figure 2.**
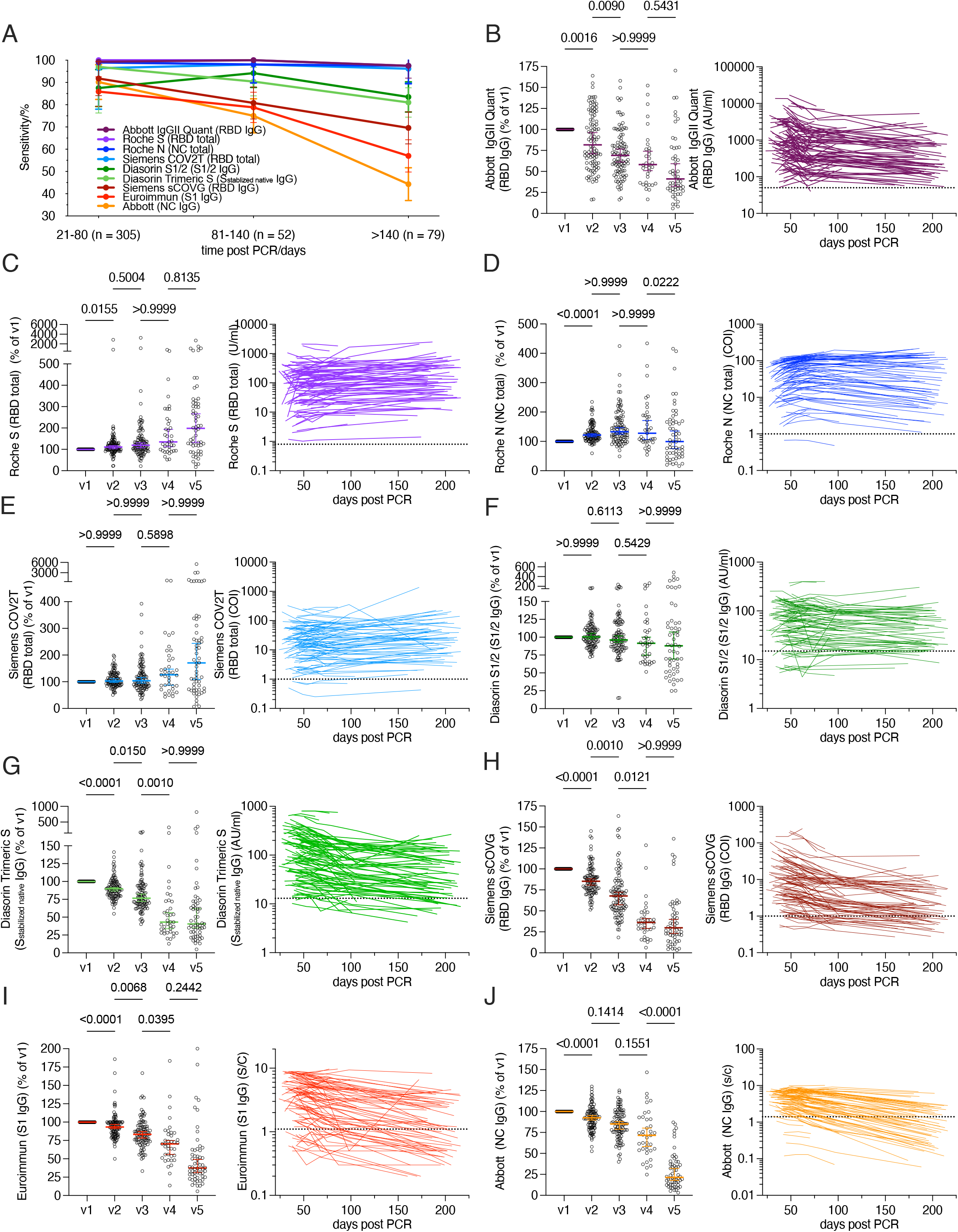
Serological analysis of longitudinal COVID-19 patient sera. **(A)** Sensitivity of the indicated serological assays in samples collected at three different time-intervals post PCR, as indicated. Mean with 95% confidence intervals are shown. **(B-J)** Relative serological results at visit 1-5 (v1-5), normalized to visit 1 (left panels) as well as serological results per participant over time with each line representing a single participant (right panels) for the assay indicated. Horizontal bars in B-J indicate median with 95% confidence interval and statistical significance was determined with the Kruskal-Wallis test and subsequent Dunn’s multiple comparisons test. Dotted lines indicate serological assay thresholds.

In addition to indicating whether a serum sample is negative or positive for antibodies against viral antigens, each assay indicates quantitative antibody levels within assay-specific scales. Analysis of antibody levels over time showed assay-dependent differences in trajectory that were not dependent on whether nucleocapsid or spike antigens were used (Figure 2A-J). Median antibody levels measured by the Roche S and Siemens COV2T assays increased slightly over time, those measured by the Roche N and Diasorin S1/S2 assays remained approximately constant, while levels measured by the Siemens sCOVG, Diasorin Trimeric S, Euroimmun and both Abbott assays decreased over time. The deviation of individual participant antibody levels from the mean increased over time in all assays except Abbott IgGII Quant which exhibited high deviations from the first time point.

We determined the ability of each serological assay to predict pseudotype virus neutralizing antibody titers over time (Figure 3, supplementary Figure 2). For each assay, the correlation with NT50 values was closest at early time points and deteriorated over time with the poorest correlation observed at visit 5 in each case. An additional assay, that measures antibodies that block the interaction between the RBD and the virus receptor (cPass), was included in this analysis using samples only from visits 1 and 5. At visit 1, antibody levels measured using the Diasorin S1/2 and trimeric S assays had the highest correlation with NT50 followed by Abbott IgGII, Siemens sCOVG, Euroimmun and cPass (Figure 3A, supplementary Figure 2). Antibody levels measured with the remaining spike antigen-based assays (Roche S and Siemens COV2T) which are designed to detect total antibody levels against the spike antigen regardless of antibody class had a lower correlation with NT50 titers, whereas the nucleocapsid based assays had the poorest correlation. For all serological assays, the correlation with neutralizing antibody titers decreased over time, but for all of the spike antigen based IgG assays (Diasorin S1/2 and trimeric S assays, Abbott IgGII, Siemens sCOVG) and cPass the correlation coefficient r, remained >0.75 even at visit 5 (Figure 3A, supplementary Figure 2).

**Figure 3.**
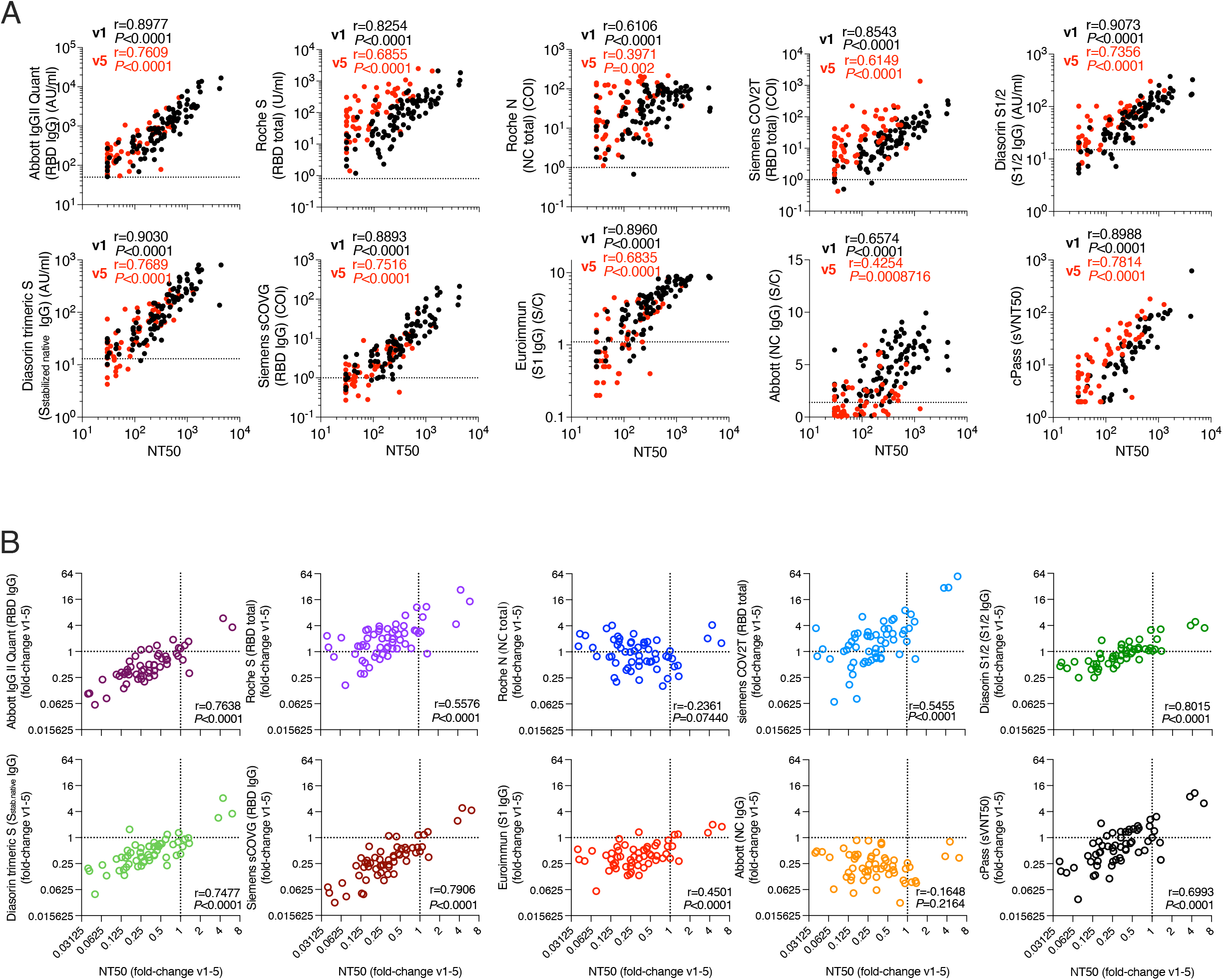
Correlation of neutralization titers and serology assays. **(A)** Correlation of NT50 (x-axis) with serological assay values (y-axis) obtained at visit 1 (black) and visit 5 (red) for each participant. Statistical significance was determined using the Spearman correlation test for samples obtained at visit 1 and visit 5 independently, as indicated. Dotted lines indicate serological assay thresholds. **(B)** Correlation of fold-change (visit1 to visit5) of NT50s with corresponding fold-change in serological assay values for indicated serology assays. Statistical significance was determined using the Spearman correlation test. Dotted lines at x=1 and y=1 indicate unchanged assay results over time.

In general, the decrease in neutralizing antibody titers over time was proportionately greater than the corresponding decrease in levels measured using serological assays (Figure 3B), particularly between early time points (supplementary Figure 2). Thus, declines in antibody levels over time measured using serological assays did not, in some cases, accurately reflect the decrease in neutralization activity. This was particularly the case for the nucleocapsid based assays, where the magnitude of the decrease in antibody measurements did not correlate with the decrease in NT50. Nevertheless, for assays that correlated best with NT50 titers at early time points, declining levels of antibodies measured in the serological assays predicted declining NT50 quite well (Figure 3B, supplementary Figure 2).

The ability of serological assays to qualitatively identify sera that did or did not have detectable neutralizing activity, was estimated (Table 1, supplementary Figure 3). This comparison was performed on sera collected across all time points and selected ‘cut off’ values for each serological assay scale were evaluated for sensitivity/specificity and predictive value (Table 1). None of the assays were effective in qualitatively discriminating neutralizing vs non-neutralizing sera using manufacturer recommended cut-offs, with specificity ranging from 3% for the Roche S and N total antibody assays to 72% for the EuroImmun assay. By selecting different cut off values the sensitivity, specificity and predictive values could be improved for some assays (Table 1).

**Table 1.**
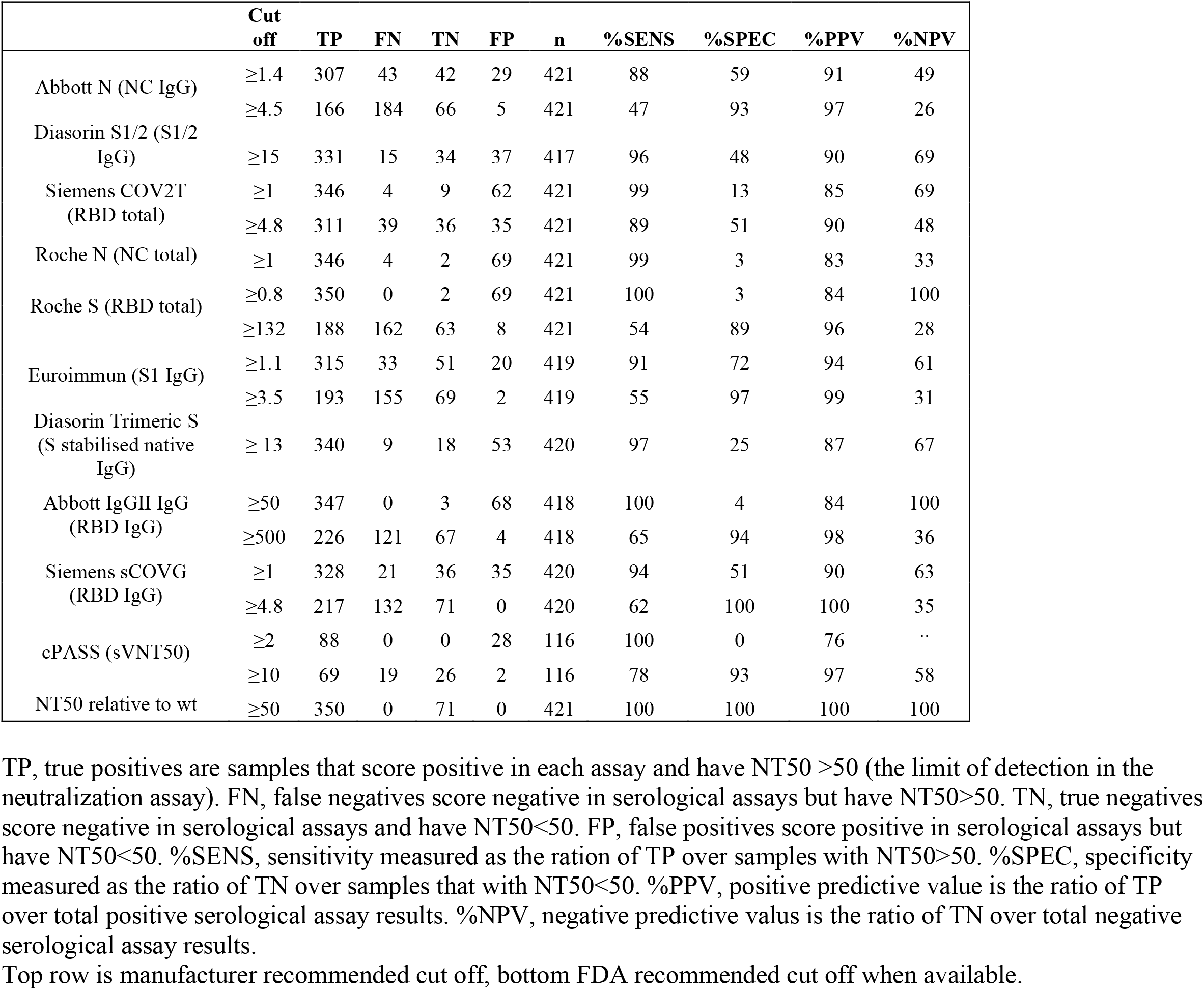
Ability of serological assays to qualitatively identify presence of neutralizing antibodies.

The occurrence of SARS-CoV-2 variants compromises the ability of first wave convalescent or vaccinee antibodies to neutralize contemporaneous viruses and might further erode the ability of serological assays based on proteins derived from the prototype (Wuhan-hu-1) virus to predict neutralizing antibody titers. We determined the ability of 58 serum samples obtained at visits 1 and 5 to neutralize selected variants of concern of the B.1.1.7, B.1.351, B.1.617.2 lineages that have been associated with partial resistance to neutralization^13-16^. Sera from both visits were able to neutralize viruses carrying the B.1.1.7 spike with potencies comparable to those observed with the Wuhan-hu-1 spike protein (Figure 4A, supplementary Figure 4). In contrast, titers against viruses with the B.1.1.7 (E484K) amino acid substitution, B.1.617.2 or either of the two B.1.351 spike variants were decreased by approximately 2.5-to-5 fold at visit 1 (Figure 4B-E, supplementary Figure 4). Differences between the ability to neutralize Wuhan-hu-1 and variant spike bearing viruses became somewhat less pronounced at visit 5 (Figure 4A-). Thus, the inclusion of amino acid substitution E484K in the context of B.1.1.7 or substitutions found in the spikes from other variants of concern appeared to significantly reduce neutralization titers, with the largest effect seen with substitutions found in the B1.351 variants, consistent with prior reports^13,14,16-20^.

**Figure 4.**
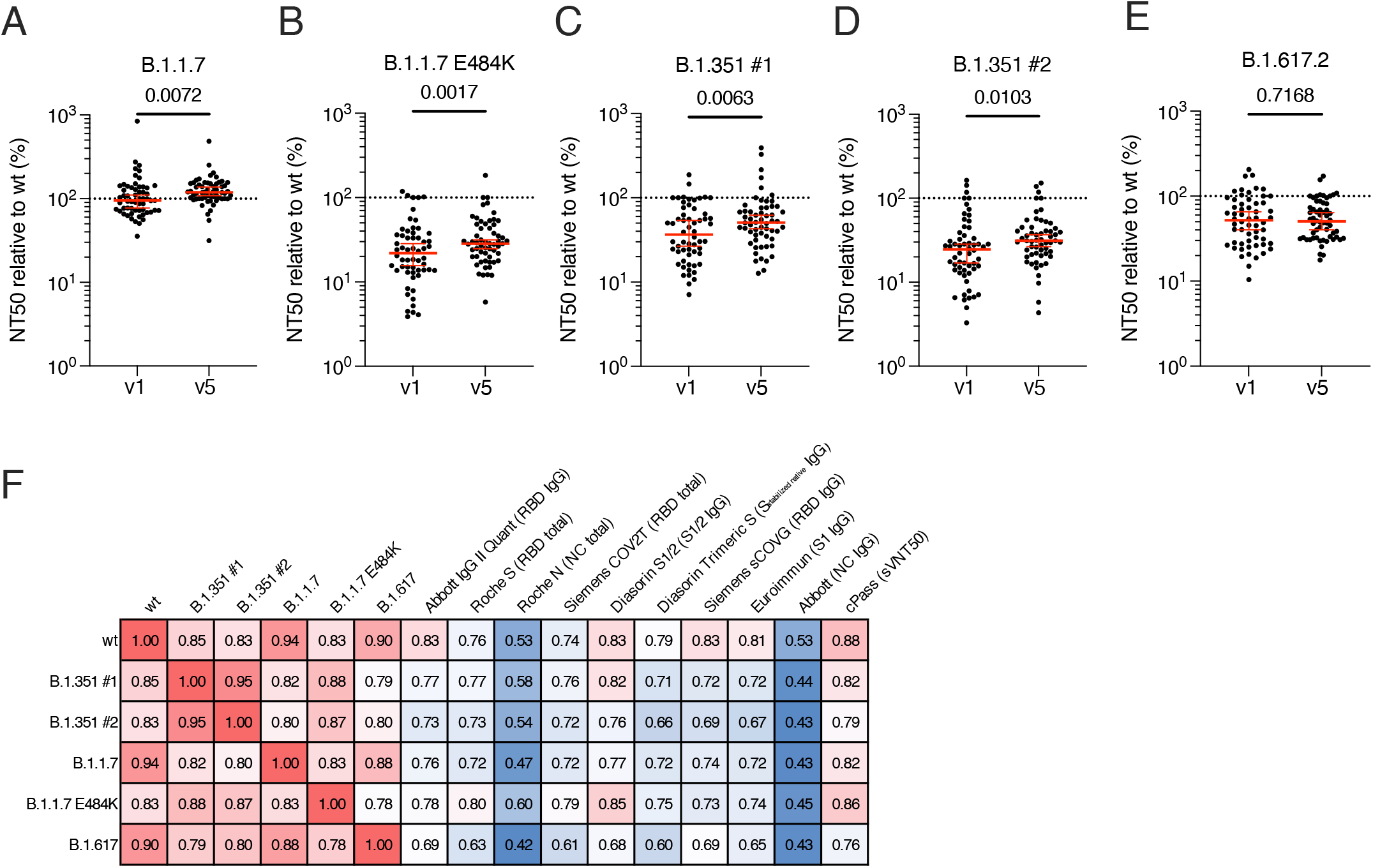
Neutralization of variants of concern. NT50, normalized to wt of pseudoviruses with spikes from variants **(A)** B.1.1.7, **(B)** .1.1.7 (E484K) **(C)** B.1.351 (#1) **(D)** B.1.351 (#2) or **(E)** B.1.617.2 measured for visits indicated. Statistical significance was determined using Wilcoxon test. Dotted line indicates 100% (equal NT50). **(F)** Correlation coefficients of NT50 values between original and variant viruses and each serological assay as indicated.

Overall, the correlation of antibody levels, as measured by serological assays, with neutralizing activity was marginally weaker for variant pseudotyped viruses than for the Wuhan-hu-1 variant (Figure 4, supplementary Figure 5). The weakest correlations were observed for the B.1.617 variant. These data suggest that some degree of variation in viruses circulating in a given population does not abolish the ability of serological assays to predict neutralizing potency against a given variant. Nevertheless, the calibration of predictions made using serological assays needs to account for which particular SARS-CoV-2 variants are in circulation in a given locale, and their susceptibility to neutralization.

## Methods

### Participants

112 Participants with a history of SARS-CoV-2 infection, diagnosed by RT-PCR were recruited^8^. Recruits were surveyed to determine the date of the positive PCR test, the date of onset of symptoms, and if their symptoms required hospitalization. Serum samples were taken at a baseline visit (∼3.5 to ∼8.5 weeks post PCR test), and 2 weeks (visit 2), 4 weeks (visit 3), 8 weeks (visit 4) and 22 weeks (visit 5) later. In total, 101 participants, completed at least 3 visits, 58 participants completed the fifth visit and were included in the neutralization assays. Four patients out of 101 were hospitalized but none required intensive care. The mean age of the participants was 44.5 years (21 – 65 y), with 72 female (71% of cohort) participants. At visit 1 (baseline), the average number of days between PCR test and visit 1 (baseline) was 41.05 days (24 – 64 days); at visit 2 (2 weeks post-baseline), the average number of days post-PCR test was 55.1 days (40 – 79 days); at visit 3 (4 weeks post-baseline), the average number of days post-PCR test was 70.06 days (55 – 95 days); at visit 4 (8 weeks post-baseline), the average number of days post-PCR test was 97.81 days (85 – 110 days); at visit 5 (28 weeks post-baseline), the average number of days post-PCR test was 194 days (160-216 days). Ethical approval was obtained for this study to be carried out through the NHS Lothian BioResource (SR1407) and London-Brent Research Ethics Committee (REC ref: 20/HRA/3764 IRAS:28653). All recruits gave written and informed consent for serial blood sample collection. De-identified samples were shipped to the Rockefeller University whose IRB reviewed and approved the study.

### Serological assays

The Abbott SARS-CoV-2 IgG (NHS Lothian) and SARS-CoV-2 IgGII (NHS Greater Glasgow and Clyde) assays are two-step chemiluminescent microparticle immunoassays (CMIA) designed to detect IgG antibodies against nucleoprotein and RBD respectively. The DiaSorin LIASON SARS-CoV-2 S1/2 IgG (NHS Lothian), DiaSorin LIASON trimericS IgG (NHS Highland) and Siemens sCOVG RBD IgG (NHS Tayside) assays are also two-step CMIA designed to detect IgG antibodies. The Euroimmun SARS-CoV-2 IgG assay (Scottish National Blood Transfusion Service) is an indirect ELISA using the S1 domain of Spike as the antigen. The Roche Anti-SARS-CoV-2 N and S assays (NHS Lanarkshire) and the Siemens COV2T assay (NHS Tayside) are two-step bridging electrochemiluminesent immunoassays (ECLIA) using nucleocapsid or the RBD of the Spike protein as antigens. The cPass assay detects antibodies that block binding of a soluble RBD to an immobilized cellular receptor protein.

### SARS-CoV-2 pseudotyped reporter virus

SARS-CoV-2 pseudotyped particles were generated as previously described^10^. Briefly, 293Tcells were transfected with pNL4-3ΔEnv-nanoluc and pSARS-CoV-2-SΔ19. 48 hours later particles were harvested, filtered and stored at -80°C.

The amino acid deletions and/or substitutions corresponding to variants of concern were incorporated into a spike expression plasmid using synthetic gene fragments (IDT) or overlap extension PCR mediated mutagenesis and Gibson assembly. Specifically, the variant-specific deletions and substitutions introduced were:

B.1.1.7: ΔH69/V70, ΔY144, N501Y, A570D, D614G, P681H, T761I, S982A, D1118H

B.1.1.7 E484K: ΔH69/V70, ΔY144, N501Y, A570D, E484K, D614G, P681H, T761I, S982A, D1118H

B.1.351 #1: L18F, D80A, D215G, Δ242-4, K417N, E484K, N501Y, D614G, A701V

B.1.351 #2: D80A, D215G, L242H, R246I, K417N, E484K, N501Y, D614G, A701V

B.1.617.2: T19R, Δ156-8, L452R, T478K, D614G, P681R, D950N

These spike proteins included the R683G substitution, which disrupts the furin cleavage site and increases particle infectivity and neutralization sensitivity. Therefore, in these neutralization assays a wildtype SARS-CoV-2 spike (NC_045512), carrying R683G was used for comparative purposes.

### Pseudotyped virus neutralization assay

Fivefold serially diluted serum from COVID-19-convalescent individuals were incubated with SARS-CoV-2 pseudotyped virus for 1 h at 37 °C. The mixture was subsequently added to 293TAce2 cl22 cells (for analyses using SARS-CoV-2 Wuhan-Hu-1 pseudovirus) or HT1080Ace2 cl14 cells (for analyses involving variant pseudovirus panels and respective Wuhan-Hu-1 R683G controls).^10^ The starting serum dilution on cells was 1:50. Nanoluc Luciferase activity in lysates was measured 48 hours post-inoculation using the Nano-Glo Luciferase Assay System (Promega) with the Glomax Navigator (Promega). Relative luminescence units were normalized to those derived from cells infected with SARS-CoV-2 pseudotyped virus in the absence of serum. The half-maximal neutralization titers for sera (NT_50_) were determined using four-parameter nonlinear regression (least squares regression method without weighting; constraints: top=1, bottom=0) (GraphPad Prism).

## Discussion

Tracking transmission dynamics, spread and prevalence of viral infections, is critical in mitigating viral epidemics, particularly when a large number of cases remain asymptomatic during infection as is the case with SARS-CoV-2. Moreover, the wide-spread use of vaccines necessitates the accurate determination of the vaccination and immune status of individuals. High-throughput serological assays address these needs but their usefulness obviously depends on their accuracy and reliability. In this study we compared the results provided by a number of SARS-CoV-2 serological assays with an emphasis on their ability to predict neutralization activity as antibodies both wane and evolve, and SARS-CoV-2 variants emerge^4,5^.

Serological assays are generally optimized for increased sensitivity so they can reliably diagnose the presence or absence of antibodies against viral antigens^1,21^. The majority of the assays used herein accomplish this goal with sera obtained shortly after infection, however, their sensitivity was not always maintained over time and certain assays exhibited a sharp decline in sensitivity at later time points after infection. This loss of sensitivity was not related to the antigen on which the assays were based.

Serological assays also provide a quantitative result that could potentially enable their use for estimation of antibody levels and prediction of immunity, especially if antibody levels correlate with functional neutralizing antibody titers. Assays that detect spike-binding antibodies can use various protein subdomains or conformations (e.g. isolated RBD, S1 or a stabilized trimeric spike) as their antigens. Moreover, some assays detect only specific antibody classes such as IgG, or those that directly interfere with RBD-receptor binding. In contrast, neutralization assays detect all antibodies capable of inhibiting spike-mediated virus entry into cells. While neutralizing antibodies are sometimes dominated by those targeting RBD^22,23^, including those that block ACE2 binding, antibodies targeting the N-terminal domain of S1 can significantly contribute to the overall serum neutralization activity in plasma^24-26^. Weak neutralizing activity has also been ascribed to antibodies targeting the region of S2 involved in fusion^27^.

Multiple studies have shown that neutralizing antibody titers following natural infection or vaccination wane over time^4,8,11,12,28^, a decline that is not always accurately reflected by serological assays. Nevertheless, certain assays used herein that detect spike-specific antibodies maintained good levels of correlation with neutralizing titers over time. Assays measuring spike-specific IgG antibodies predicted neutralizing antibody titers more accurately than those measuring total antibodies against spike or those against the nucleocapsid protein. The Diasorin assays, Abbott IgGII, Siemens sCOVG, Euroimmun and cPass had the highest correlation with neutralizing antibody titers across all comparisons and changes in quantitative values over time for these assays were most closely correlated with changes in neutralizing antibody levels within individuals. The quantitative results from these assays are therefore best suited for estimating neutralizing antibody levels at a population level. In contrast, the qualitative assay results which, if based solely on the manufacturer recommended cut-offs, are poorly specific for detecting the presence of neutralizing antibodies and may lead to a significant over-estimation of antibody related immunity. For some assays it may be possible to improve specificity for the presence of neutralizing antibodies through selecting a higher quantitative cut-off value, thus improving the positive predictive value of these assays for neutralizing antibody detection.

The majority of the naturally infected population studied thus far were infected with viral variants closely related to a prototype variant (Wuhan hu-1). The antigens used by all serological assays rely on protein sequences derived from that prototype. However, over the last several months new variants have emerged that encode multiple amino acid substitutions in their spike proteins, some of which impact neutralization by convalescent or vaccinee antibodies^13,14,16,17,29,30^. Our data indicate that antibody levels measured using several serological assays maintain good correlation with neutralization titers against some of the most important variants that have emerged thus far. However, this property will need to be monitored in the future if serological assays are to be used to predict immunity, particularly as antibodies diversify in response to variant virus infection and, potentially, variant booster vaccination.

The need for serological assays in monitoring natural infection at the population level remains. Moreover, the introduction of vaccines raises new requirements for serological assays. These requirements include (i) distinction between vaccinated and naturally infected individuals, (ii) prognostication of levels of protection against infection and disease afforded by vaccines and (iii) identification of individuals where a boosting immunization or monoclonal antibody therapy is indicated. Furthermore, in instances where countries are considering deploying ‘immunity passports’ to allow, for example travel, the selection of assays and the diagnostic cut off used can have important ramifications. Finally, since measurement of antibody function (e.g., neutralization) at a population level is not currently practical, establishing the ability of serological assays to predict neutralization and immunity will help determine correlates of protection that can be applied at a large scale, and perhaps as part of routine clinical practice.

## Data Availability

De-identified data will be available for share following manuscript publication.

## Contributors

HW, SJ, TH and PDB conceived and designed the study. FM performed and analyzed the neutralization and cPASS assays and performed cross-assay data analyses and data visualization. HWKT and SJ acquired and analyzed data using the serological assay platforms. HW and SJ performed ROC and sensitivity analyses and respective visualization. HW, SJ and MS provided Abbott N data. EF provided Siemens data. SM provided DiaSorin data. IG and AB provided Abbott S IgG II data. LJ and KM provided EuroImmun data. CR and JM provided Roche data. BB and KM assisted with collection of research clinic samples. FZ and FS provided spike plasmids. TH wrote the manuscript with help from FM, PDB, SJ and input from all authors. All authors had access to all the data in the study and accept responsibility for publication.

## Acknowledgements

We acknowledge the support of NRS BioResource and NHS Lothian Outpatients with the provision of this research study. We are grateful to Dr Linfa Wang and GenScript for providing the cPass assay kits.

Funding sources NIH (R01AI78788 to TH, R01 AI50111 to PDB) and NRS BioResource (SR1407 to SJ).

## Data sharing

De-identified data will be available for share following manuscript publication.

## Figure Legends

**Supplementary Figure 1.**
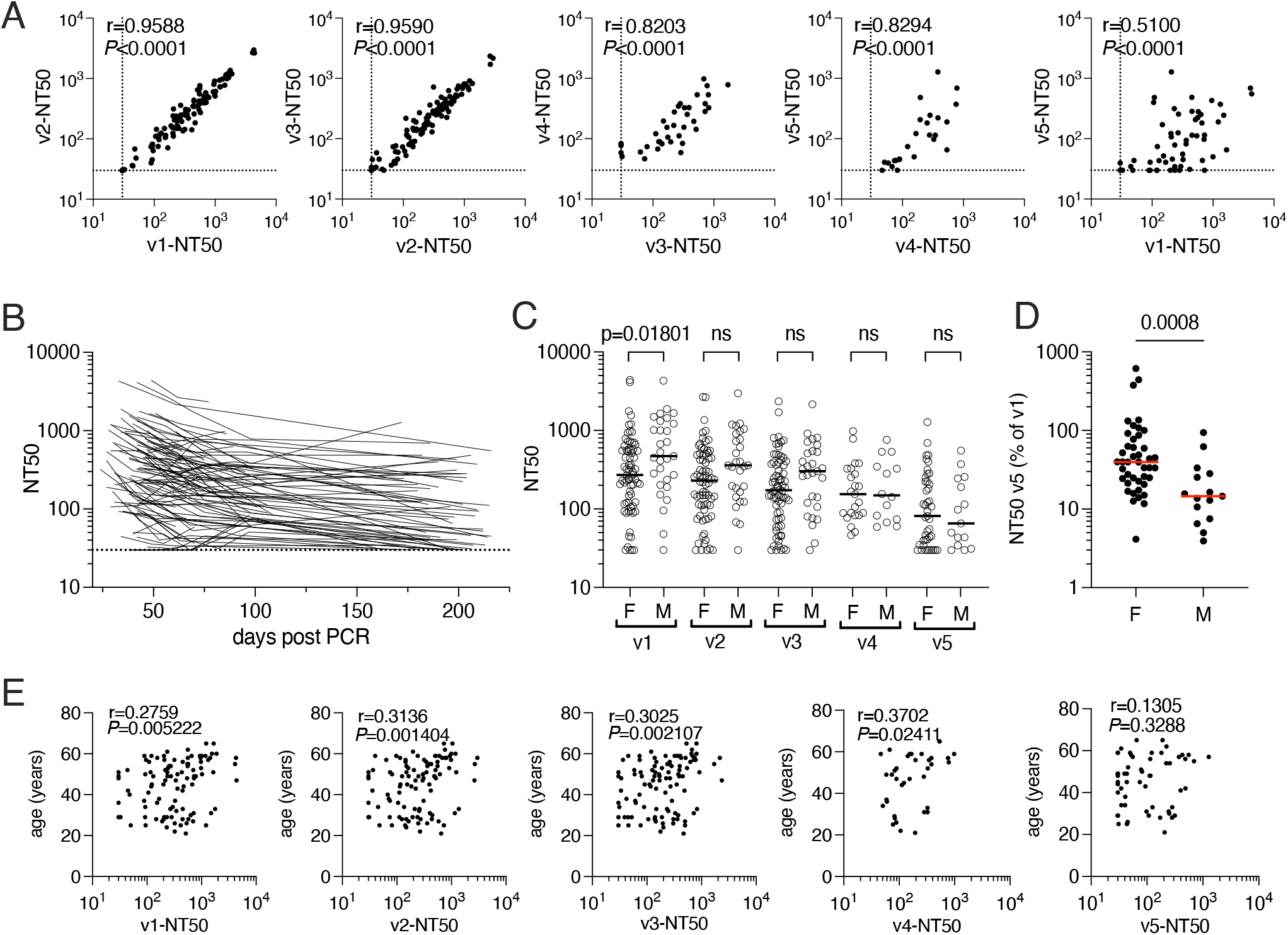
Correlation of neutralization activity and demographic parameters. **(A)** Correlation of NT50s in samples collected at different timepoints. **(B)** NT50 values per patient over time with each line representing a single participant. **(C)** NT50 values in female and male participants, sampled at the indicated visit. **(D)** Relative NT50 values at visit 5, normalized to visit 1 for female and male participants. Statistical significance was assessed with the Mann-Whitney test. **(E)** Correlation of NT50 and age at the indicated timepoints. Statistical significance in (A) and (E) was determined using the Spearman correlation test. Dotted lines in (A) and (B) indicate limit of detection.

**Supplementary Figure 2.**
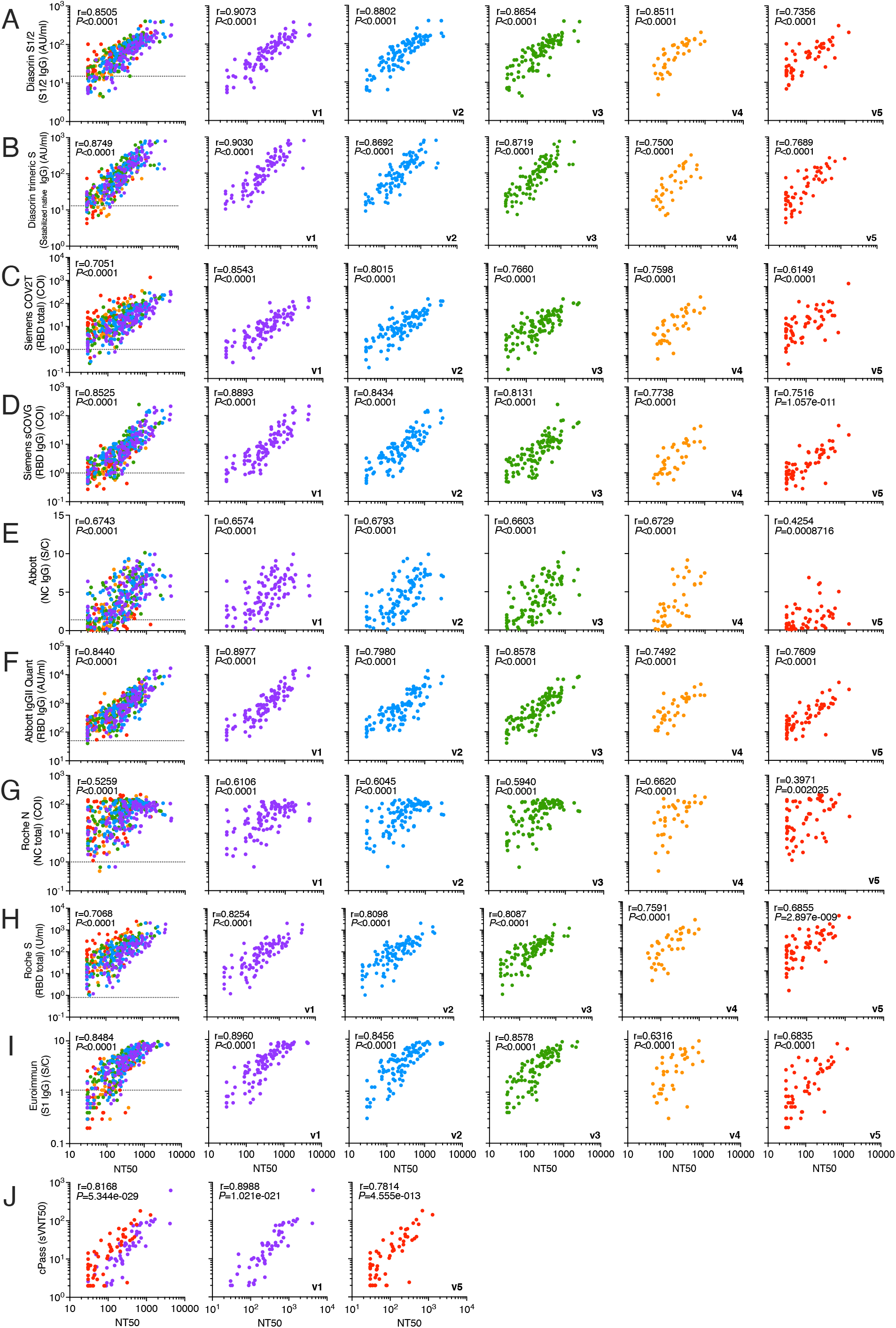
Correlation of neutralization titers and serology assays per visit. **(A-J)** Correlation of NT50 (x-axis) and indicated serological assay measurements (y axis) at visit 1 through 5. Sampling timepoints are indicated by color and shown collectively (left) and individually. Statistical significance was determined using the Spearman correlation test and r and *P*-values are indicated for total samples (left panels) and for the individual visits. Dotted lines in left panels indicate serological assay thresholds.

**Supplementary Figure 3.**
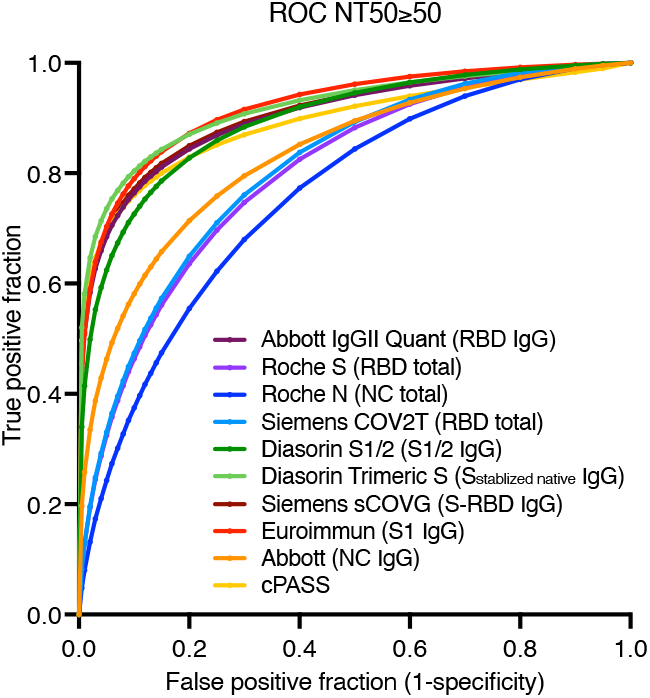
ROC analysis. Receiver-operating characteristic curve (ROC) for prediction of NT50>50 based on results obtained with indicated serology assays. Shown are mean values.

**Supplementary Figure 4.**
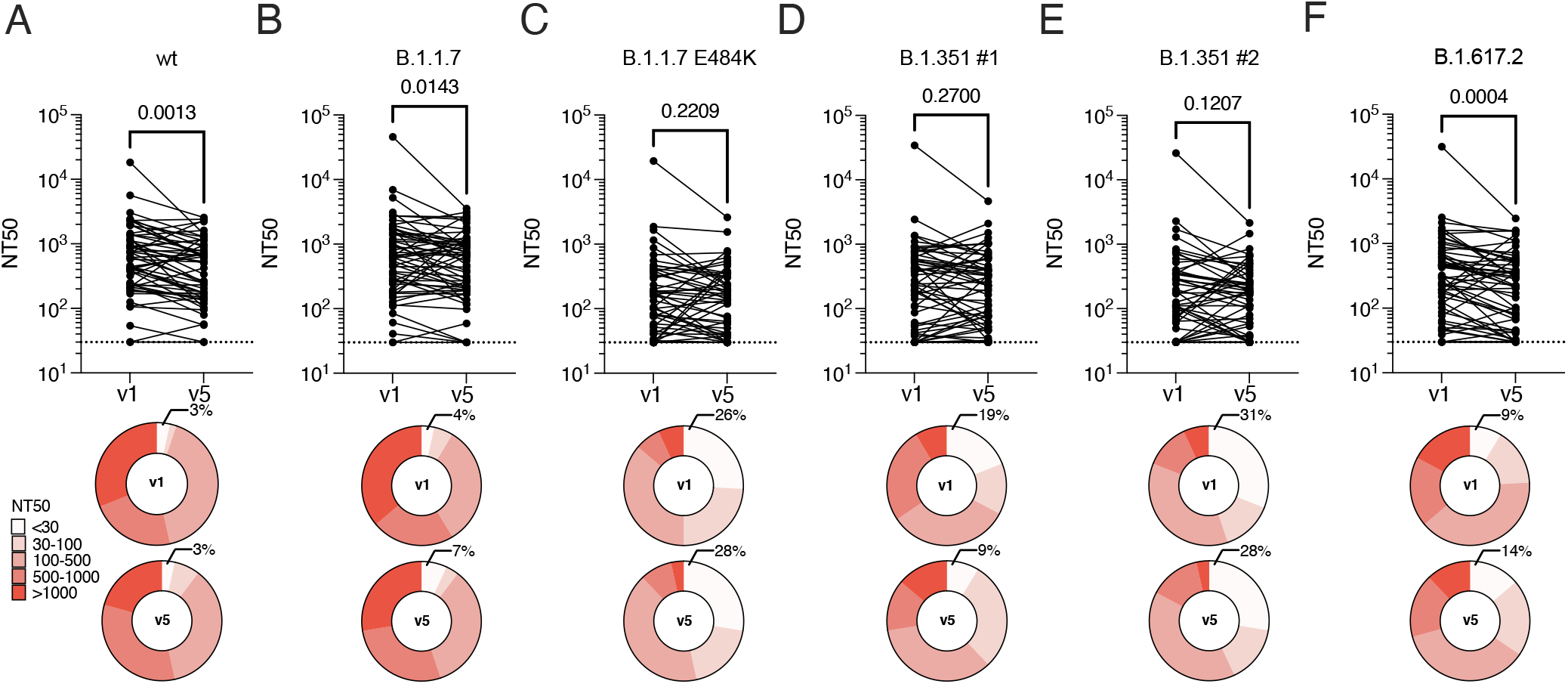
Neutralization of variants of concern. NT50s for wt **(A)** B.1.351 (#1) **(B)**, B.1.351 (#2) **(C)**, B.1.1.7 **(D)** and B.1.1.7 (E484K) **(E)** pseudovirus at visit 1 and visit 5. Statistical significance was determined using Wilcoxon test. Dotted line indicates limit of detection. Deletions/substitutions present in VOCs, as well as respective wt control are in R683G background, as indicated.

**Supplementary Figure 5.**
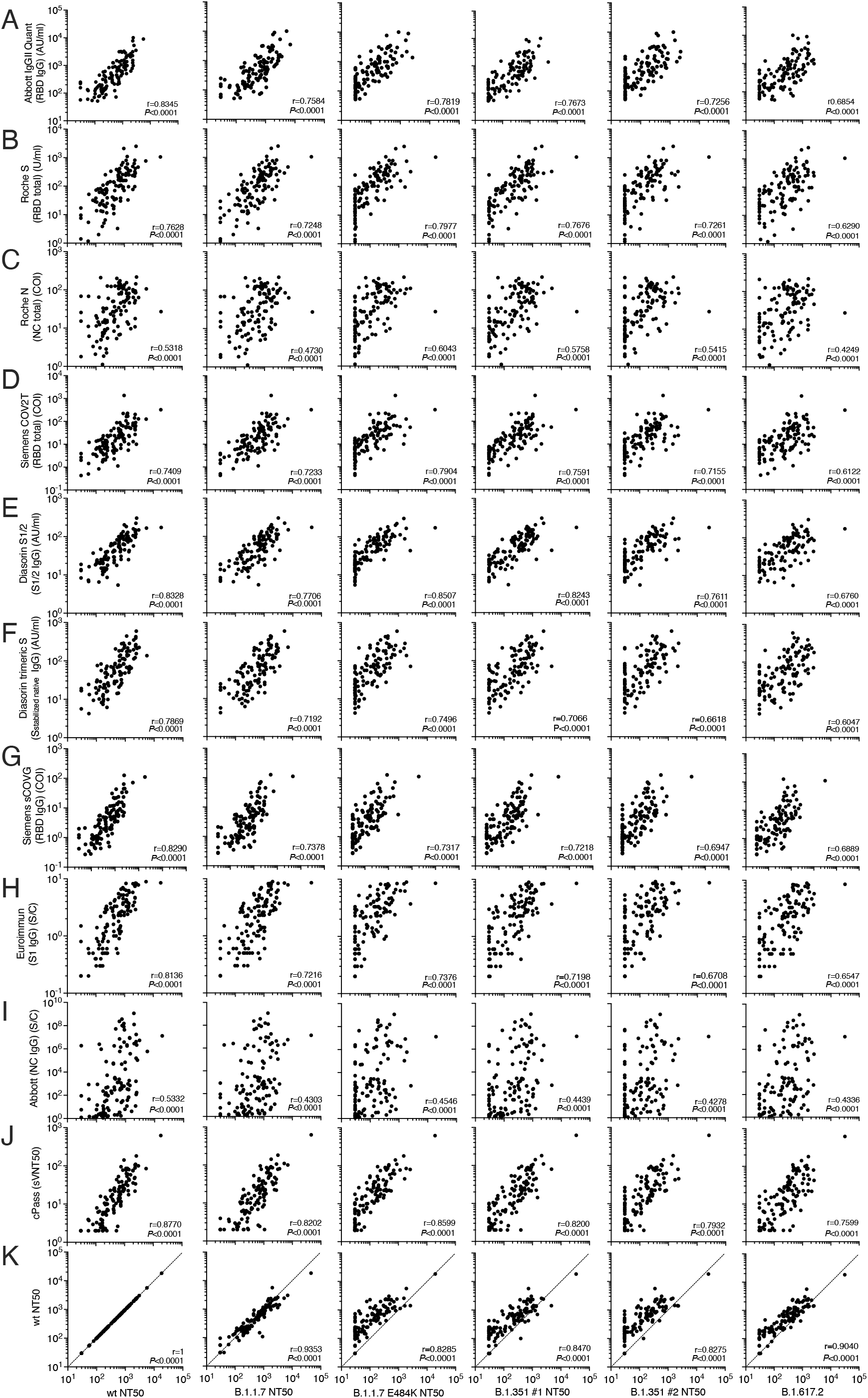
Neutralization of variants of concern versus serology assays. **(A-K)** Correlation of, from left to right NT50s against the pseudoviruses indicated with the Abbott IgGII Quant **(A)**, Roche S **(B)**, Roche N **(C)**, Siemens COV2T **(D)**, Diasorin S1/2 **(E)**, Diasorin Trimeric S **(F)**, Siemens sCOVG **(G)**, Euroimmun **(H)**, Abbott NC **(I)** and the cPass assay **(J)** as well as with wt NT50 values. Samples obtained at visit 1 and visit 5 are included. Statistical significance was determined using the Spearman correlation. Deletions/substitutions present in VOCs, as well as respective wt control are in R683G background.

